# Global comparison of influenza A and B epidemiology identifies consistent geographic and socio-demographic predictors

**DOI:** 10.64898/2026.03.14.26348363

**Authors:** Christian E. Gunning, Shirin Rezaeimalek, Pejman Rohani

## Abstract

Seasonal influenza outbreaks are caused by types A and B that together account for an estimated 3-5 million severe cases each year. Most attention has focused on influenza A viruses (IAVs) due to their rapid evolutionary dynamics and high disease burden, and has been concentrated in well-observed high-income regions. Here, we use a macroecological approach to compare and contrast the global epidemiology of IAVs and influenza B viruses (IBVs) across 111 countries and 15 influenza seasons (2010-2024). We first show how temporal correlations between countries depends on both distance and geographic region. For both IAV and IBV, we find high overall synchrony among northern temperate countries, whereas tropical countries display marked heterogeneity. At the longer time scale of influenza seasons, we next quantify sampling intensity, positivity, seasonality, fade-out dynamics and the timing and variability of epidemic peaks. We then describe how these long-term epidemiological outcomes change in association with a suite of 17 geographic, climatic, and socio-economic variables. In addition, we document persistent surveillance gaps, particularly in Africa, and highlight ongoing but spatially variable impacts of the SARS-CoV-2 pandemic-era on sampling. Overall, we find strong correspondence between the macroscopic features of IAV and IBV epidemiology, with critical roles played by geography and climate (especially latitude and temperature), economics (*per capita* GDP) and demographics (population size and *per capita* birth rate).

**Significance Statement:** The global circulation of seasonal influenza A and B viruses (IAV and IBV) imposes major human health impacts each year that very widely across space and time. An improved understanding of these dynamics could improve public health preparedness, response, and intervention efforts. Here we offer a comprehensive comparison of IAV and IBV dynamics across 15 seasons, 111 countries, and six continents. We demonstrate the impact of distance and region on temporal correlation, quantify how measures of influenza seasonality change with geographic and socioeconomic factors, and predict how frequently influenza cases are absent from countries. Our study finds widespread similarities between IAV and IBV (along with key differences), documents notable geographic clusters of countries with shared dynamics, and highlights persistent gaps in global influenza surveillance.

---

**S**easonal influenza poses a major public health challenge, affecting an estimated 1 billion people worldwide each year. Of these cases, 3 to 5 million cases are classified as severe, leading to approximately 290,000 to 650,000 respiratory-related deaths (1). This burden is primarily linked to two types: influenza A viruses (IAVs) and influenza B viruses (IBVs) (2, 3). IAVs are typically more widespread and cause greater mortality (4–6) as well as numerous high-impact global pandemics (7, 8). In contrast, IBVs are generally less virulent and cause lower morbidity, and have consequently received far less attention (9–11). Despite their recognized difference in disease severity, however, both IAV and IBV continue to impose substantial global disease burdens, particularly in vulnerable populations (12–14).

Both IAV and IBV evolve rapidly and spread globally, leading to characteristic seasonal epidemics that evade existing population immunity and vaccination efforts (15–17). This has led to an intense focus on identifying those factors that facilitate the global transmission of influenza (18–20), as well as those factors shaping more localized dynamics (21–23). In addition, the human impacts of these viruses also vary widely, with demographic and socioeconomic factors playing a critical role (24–26). An improved understanding of such factors could aid public health preparedness and responsiveness at both local and global scales, yet a unified view of influenza dynamics remains elusive. This issue has been further complicated by low and variable disease surveillance, particularly in low- and middle-income countries (27–29), along with widespread correlations between many of the climatic, demographic and socioeconomic factors thought to shape influenza dynamics (and surveillance efforts) (30–32).

While many high-income countries have systematic influenza tracking systems in place, much of what we know about influenza at global scales relies on the World Health Organization’s (WHO) FluNet program (33). Established in 1997 under the Global Influenza Surveillance and Response System (GISRS), FluNet has become a keystone of international influenza monitoring, enabling near real-time reporting of laboratory-confirmed cases from over 180 countries (34). It compiles weekly counts of specimens tested and confirmed influenza cases by influenza type (and, less frequently, subtype). The program employs standardized protocols (35) that are followed by National Influenza Centres and collaborating laboratories. Since its launch, FluNet has expanded dramatically: National Influenza Centres testing some 3.4 million clinical specimens annually between 2014 and 2019, a figure that surged to 6.7 million influenza tests during 2020-2021 (36). Despite persistent disparities in surveillance capacity and under-reporting in low-resource settings (29, 37), FluNet remains a critical global resource, supporting influenza research, vaccine strain selection, and pandemic preparedness (38–40).

The temporal dynamics of seasonal influenza viruses are recognized to vary widely across the world’s regions and countries (26, 41). Most notably, temperate regions typically experience pronounced seasonal outbreaks, while tropical regions tend to have more variable, year-round activity (42–46), with humidity and temperature identified as likely drivers in shaping these temporal dynamics (29, 47–49). And while most of this work has focused on IAV, several comparative global studies have examined the influence of climate on both IAV and IBV (29, 50).

Critically, influenza’s marked seasonality contributes to recurring local elimination and reintroduction that is thought to play a central role in shaping influenza evolution (3, 16, 20, 51). Previous work has identified human mobility, particularly air travel, as a central driver in facilitating the spread of influenza within and between regions (18, 20, 23, 52).

Here we provide a comprehensive global synthesis of seasonal influenza epidemiology. We adopt a data-driven, macroecological approach (53) in order to A) characterize patterns of IAV and IBV epidemiology across a broad range of space and time and B) identify potential drivers of these patterns. Employing countries as our basic unit of measure, we first explore the spatial structure of temporal correlations among countries. We then quantify the seasonal epidemiology of IAV and IBV within each country, including testing intensity, positivity, absence of case reports, and several measures of seasonality. We match these outcomes against a comprehensive set of demographic, socioeconomic, epidemiological, and climatic predictors. We also document major shifts in global influenza surveillance during and after the 2020 SARS-CoV-2 pandemic. Our analysis provides a nuanced view of influenza dynamics both within and among countries, particularly in areas where surveillance systems have traditionally been lacking, along with a direct comparison between IAV and IBV epidemiology.

Given that the timing and geographic distribution of influenza activity have direct implications for public health preparedness, our study offers critical insights into the broader epidemiological and ecological mechanisms governing seasonal influenza transmission while exposing shared features between IAV and IBV. By identifying conserved patterns of influenza dynamics and transmission, our findings can inform surveillance strategies, enhance pandemic preparedness, and aid in the development of targeted surveillance and intervention measures tailored to the distinct characteristics of each location and virus type.

## Results

Historically, influenza surveillance has been concentrated in high-income countries of the Northern temperate (N Temp) region, where widespread, synchronized seasonal IAV outbreaks have long been observed. However, the synchronized seasonality of N Temp countries is largely lacking in the tropics (42), whereas Southern temperate (S Temp) countries are generally synchronized and out of phase with N Temp countries (54). In Figure 1A, we illustrate these patterns with IAV case reports (aggregated by month) from three populous countries in each of the above regions (N Temp, Tropics, and S Temp). We also include monthly positivity (*i*.*e*. cases/tested samples) of IAV and IBV averaged across each region (Fig. 1B). In both cases, a sharp drop in global influenza incidence is apparent immediately after the onset of the 2020 SARS-CoV-2 pandemic (55–57). Notably, after controlling for testing intensity (Fig. 1), positivity records indicate that apparent post-pandemic spikes in IAV incidence (.e.g, Fig.1A) is likely a product of increased surveillance and changes in diagnostic methods (such as multiplex assays (58, 59)).

**Fig. 1.**
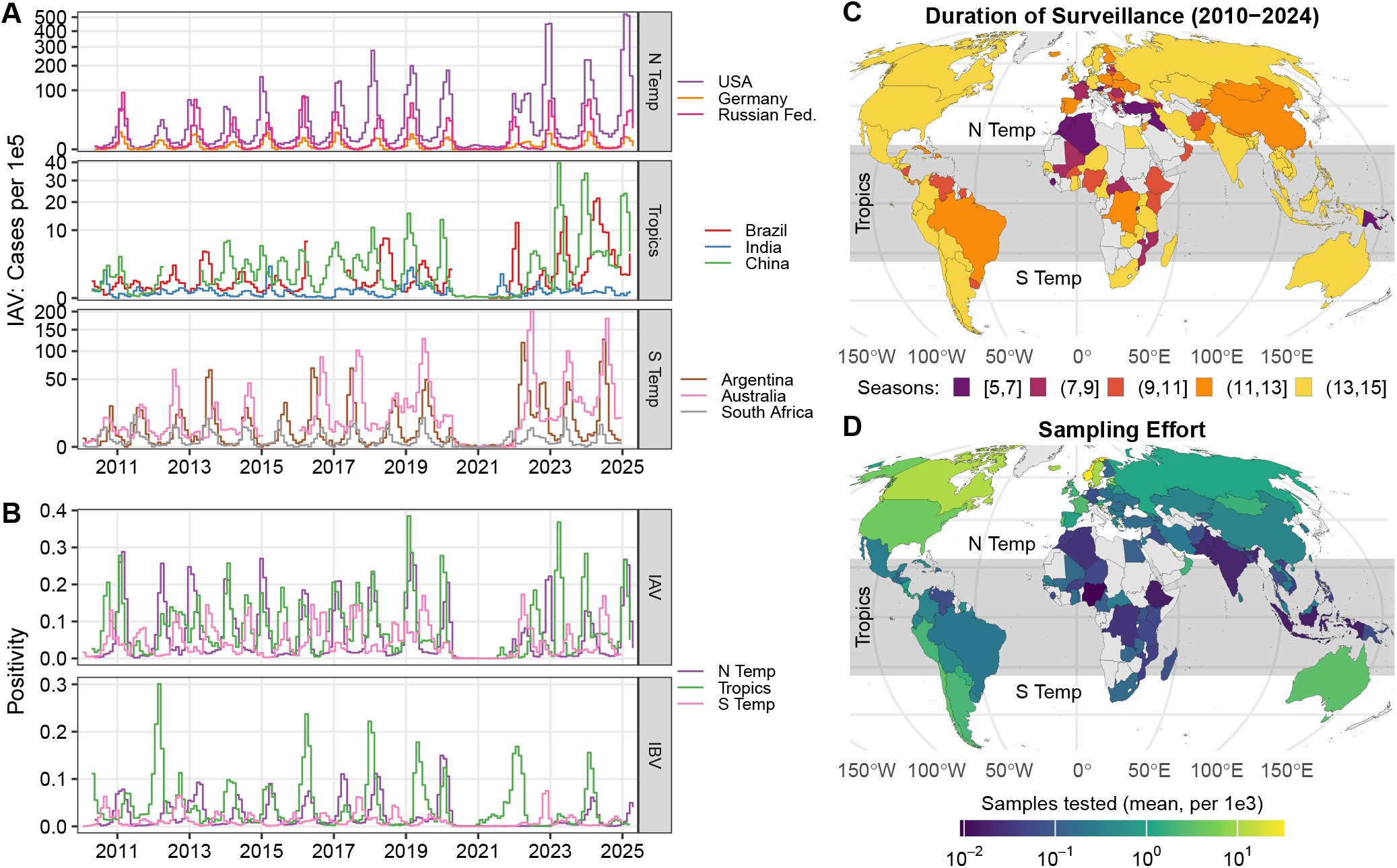
Overview of global influenza surveillance. A) Timeseries of IAV case reports (monthly, *per capita*) for select countries (color) by region (row). Synchronized seasonality is evident in the Northern and Southern temperate regions, but largely absent in the tropics. B) Timeseries of positivity (cases/tested samples, monthly) of IAV and IBV (rows) averaged across region (color). A sharp and widespread drop in positivity is evident during the SARS-CoV-2 pandemic that starting in 2020. However, once testing intensity is accounted for, we find no evidence of a widespread post-pandemic “surge” (though comparisons with the pre-pandemic era are complicated by a sharp uptick in testing volume – see Figure S3). C-D) Geographic overview of C) surveillance duration and D) mean per capita testing intensity of 111 countries (N Temp=42, Tropics=60, S Temp=9, 1,341 country-seasons). Some high-income countries were excluded due to lack of sample testing reports (e.g., Japan), whereas many low-income countries did not meet the minimum surveillance threshold of at least 5 seasons of 9 months each (pre-2020, minimum 30 weeks per season and 20 IAV cases per year). Surveillance coverage is absent across much of Africa (C), while low testing intensity (D) is evident in many tropical countries (D). Nonetheless, coverage across the full period of record is available for most of the world’s population.

Influenza surveillance duration and intensity varies widely between countries, as illustrated in Figure 1C-D. While FluNet surveillance began in 1997, country participation was initially limited to a few higher-income countries. The 2008/9 H1N1 pandemic saw a dramatic expansion in FluNet, both in participating countries and in (*per capita*) samples tested within each country. To facilitate comparisons among countries, we focus here on the 2010 through 2024 influenza seasons. Subsequent to 2010 we observe some increases in testing intensity over time, particularly post-2020, but these differences are modest relative to the more than 100-fold difference in mean testing intensity among countries (see Fig. S3-S4). We include all countries with at least 5 seasons of pre-2020 surveillance (minimum 9 months per season) for a total of 111 countries (N Temp=42, Tropics=60, S Temp=9) across 1,341 country-seasons. For reference, we provide time series of monthly tests and positivity for all included countries (Fig.S1-S2).

### Spatial determinants of temporal synchrony

In order to quantify temporal synchrony of seasonal influenza among countries within and between regions, we computed Spearman correlation (*ρ*) of monthly incidence between all pairs of countries for both IAV and IBV. In Figure 2A, we illustrate these correlations for IAV between the 50 most populous countries in our dataset, where each country is colored by region. In addition, countries are ordered by hierarchical clustering of similarity, shown as a dendrogram on the left margin (see Figure S8 for all countries). Our results demonstrate a consistent positive correlation among N Temp countries, most of which form a single large cluster. We observe weaker and more variable correlations between tropical countries, which fall into several distinct clusters, indicating heterogeneous dynamics among these countries. We also observe consistent negative correlation between S Temp and most N Temp countries (60). A similar but weaker pattern is observed with IBV (Fig. S7-S9), where N Temp and S Temp countries cluster together less consistently.

**Fig. 2.**
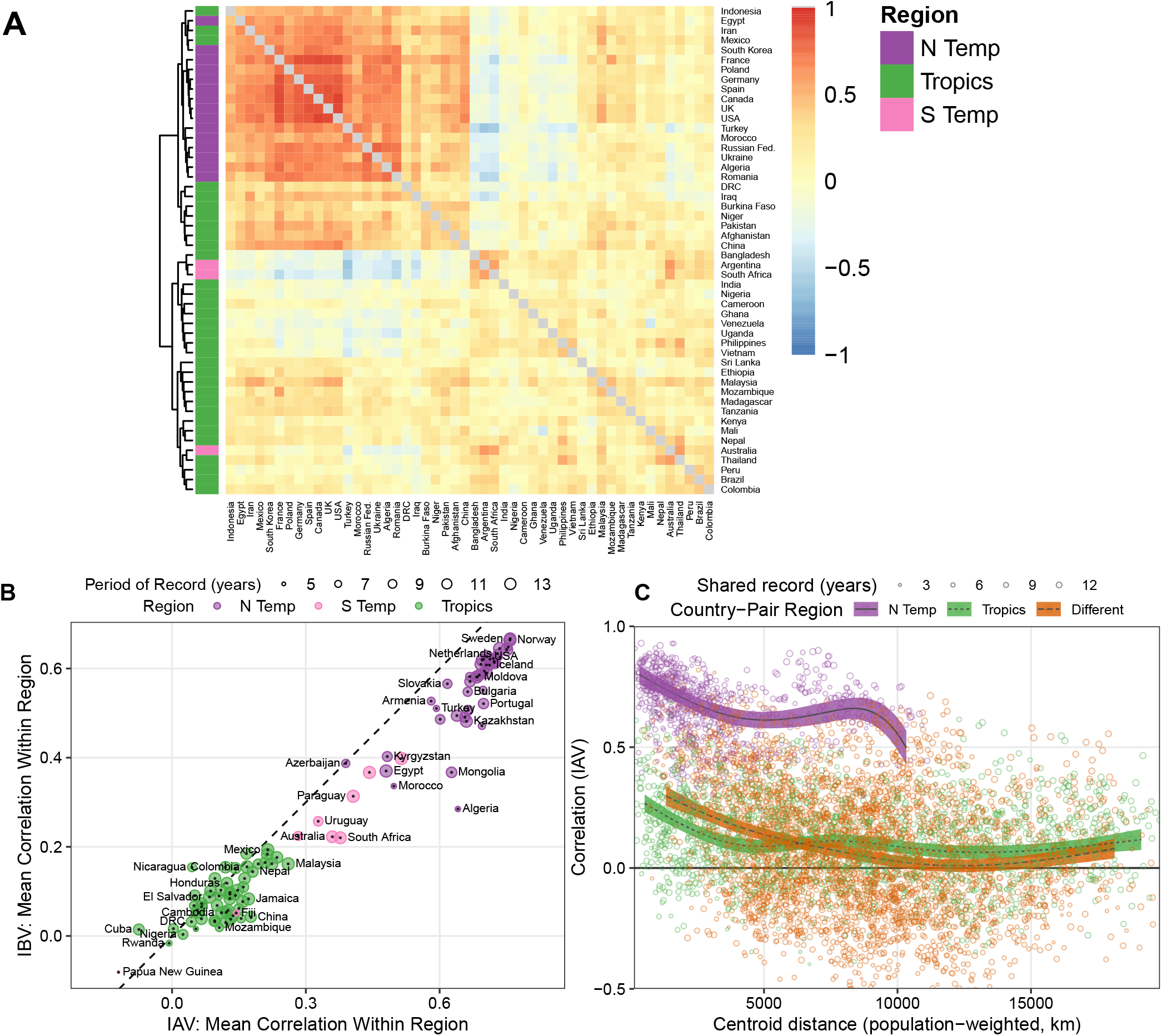
Temporal correlation of weekly IAV case reports by region and distance. A) Heatmap of Spearman correlation (*ρ*) between country pairs. Rows are clustered (and ordered) by similarity and color-coded by region (left border), which reveals consistently strong correlations between N temperate countries and wide variation among tropical countries. C) Per-country mean *ρ* between countries within each region (color) for IAV versus IBV (X vs Y; dashed black line shows X=Y). The low overall correlation between tropical countries is evident. For most countries, mean *ρ* is higher for IAV than IBV (X¿Y). C) Model results showing dependence of IAV *ρ* on between-country distance (X) and region (color; points showing individual country pairs). Among N Temp country-pairs, correlation starts high and drops modestly with distance. Among tropical and discordant-region country-pairs, correlation starts much lower and exhibits a modest decrease with distance. We find similar patterns for IBV (see Figure S11). Ribbons show model-estimated means plus 95% CI from a random effects GAM. Detailed heatmaps of showing correlation among all country-pairs, see Figures S7-S9.

To better illustrate geographic differences, we summarize the above temporal correlations by country. Figure 2B shows the median correlation of each country with other countries within the same region (color) for both IAV and IBV. Here we find a close correspondence between IAV and IBV, though IAV correlations are generally higher. Correlation among tropical countries is uniformly low, whereas N Temp countries are more correlated but more variable.

Temporal correlation by distance has been previously described for a range of infectious diseases (61–65), though the strong seasonality of influenza adds a potential layer of complication. In order to disentangle the relative impacts of region and spatial proximity, we construct a pair of generalized additive models (GAM, one model per influenza type) that predict temporal correlation between country-pairs. These models use distance (as measured from population centroids) as smooth effects (within each region) and country identities as random effects. In Figure 2C we present model fits for IAV (lines show estimated marginal means with 95% CI) along with the observed correlations (one point per country-pair). We find that temporal correlation starts high and drops modestly with distance among N Temp country-pairs, while correlation starts lower and drops modestly among tropical and discordant-region country-pairs. We find comparable results for IBV, albeit with modestly lower correlations for the closest N Temp and discordant country pairs (Fig. S11). In addition, these GAMs offer robust fits of observed correlations, with adjusted *R*^2^ of 0.68 (IAV) and 0.67 (IBV).

### Predictors of seasonal epidemiology

Having identified spatial determinants of temporal correlations, we next examine a set of epidemiological fingerprints at the seasonal scale (23, 47, 49). In Figure 3A-E, we present maps for five seasonal IAV outcomes (averaged across available seasons): A) positivity, B) seasonality, as measured by the intra-season coefficient of variation (CV) of case reports, C) percent zero weeks (fade-out dynamics, *i*.*e*., percent of weeks per year with zero cases), D) peak month and E) inter-season variation in peak timing, as measured by the (circular) maximum absolute deviation (MD) of peak month. Finally, in Figure 3F, we show the mean difference in peak timing between IAV and IBV. In addition, a direct comparison between IAV and IBV is provided in Figure S12.

**Fig. 3.**
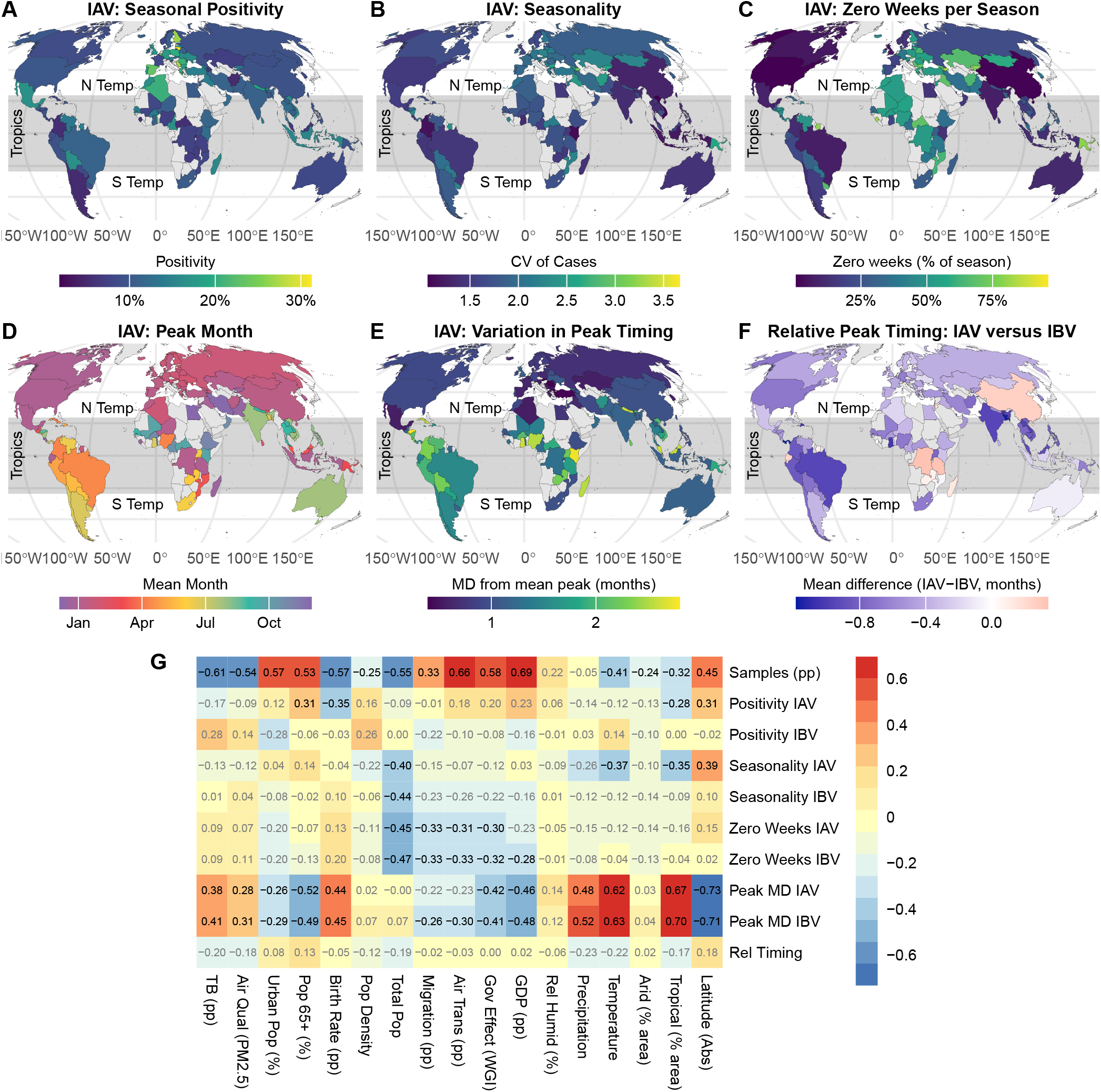
A-E) Geographic distribution of mean epidemiological outcomes for IAV by country (averaged across all seasons), including A) positivity, B) seasonality: coefficient of variation (CV) of cases, C) percent zero weeks, D) peak month, and E) inter-season variation in peak timing: maximum absolute deviation (MD). F) Mean difference in peak timing between IAV and IBV. G) Pairwise Spearman correlation (*ρ*) between each outcome (rows) and putative predictor (columns, per-country medians). Numbers show *ρ*; columns are clustered and ordered by similarity (shown in top dendrogram). Maps that offer direct comparisons between IAV and IBV are shown in Figure S12. Correlations among outcomes and among predictors are shown in Figures S13-S14.

For IAV outcomes, we observe noteworthy spatial clustering, pointing to the impact of geography, climate, and/or socioeconomic conditions. For IAV positivity (Fig. 3A), the top five countries are all European: Lithuania (31%), Serbia (28%), Finland (27%), Belgium (25%), and Spain (23%). For seasonality, three out of the top five countries are in the Black Sea Region and are separated by less than 1,500 km, including Azerbaijan (1st, CV=3), Bulgaria (4th, 2.6) and Armenia (5th, 2.5), along with the Central Asian countries of Kyrgyzstan (3rd, 2.6), Afghanistan (6th, 2.5), and Iraq (7th, 2.5). Peak timing (Fig. 3D) of most N Temp countries is tightly synchronized to late winter (Jan-Feb) whereas South American countries notably cluster around early summer (May). The most consistent (least variable) peak timing (Fig. 3E) is found in Romania (0.39 months), Slovakia (0.4), Turkey (0.41), Bulgaria (0.48), and Algeria (0.56), four of which are Eastern European countries. In most cases, the peak of IAV leads that of IBV (Fig. 3F; 99/111), while most exceptions occur within tropical countries.

We systematically evaluate the relationship between each of the above outcomes with a comprehensive suite of 17 putative predictors, including geographic, climatic, socioeconomic, and demographic factors. In Figure 3G, we display the pairwise Spearman correlation between each outcome (row) and predictor (column), and indicate statistically significant relations in bold (using a false discover rate adjustment with *α* = 0.05 (66)). *Per capita* (pp) testing volume is strongly predicted by pp GDP (*ρ* = 0.69). Absolute latitude is strongly associated with several outcomes, notably peak variability (peak MD) of IAV (*ρ* = −0.73) and IBV (*ρ* = −0.71), while population size is associated with seasonality of IAV (*ρ* = −0.40) and IBV (*ρ* = −0.44), and with zero weeks of IAV (*ρ* = −0.45) and IBV (*ρ* = −0.47). Of note, we observe similar patterns of correlation between IAV and IBV for zero weeks and peak MD but not for positivity or seasonality. Finally, several outcomes are only weakly associated with the tested predictors, including IBV positivity and the relative peak timing of IAV and IBV.

We note that percent zero weeks of both IAV and IBV is modestly correlated with several factors, including population size, air travel, and government effectiveness. Upon closer inspection, we find that a simple linear model predicting IAV zero weeks from region, population, and government effectiveness has an adjusted *R*^2^ = 0.56 (DF=105, largest *p <* 0.005) and, for IBV, *R*^2^ = 0.51 (DF=105, largest *p <* 0.01). Whether these observed zero weeks (which includes only reported zeros and not missing reports) represent true interruption of disease transmission (versus an absence of *evidence* of incidence) is unclear. Nonetheless, the observed patterns are consistent with classic metapopulation theory, which suggests that smaller and more isolated populations will experience more frequent local extinction (67–72).

For context, we quantify the correlation structure among each pair of predictors (Fig. S13). Consistent with previous work (23, 49, 73–75), we find distinct blocks of correlated (and anti-correlated) predictors that cluster broadly into indicators of wealth (*e*.*g*., GDP, Pop 65+, and air transit), and of poverty (*e*.*g*., temperature, birth rate, and TB burden). We also note substantial differences in reporting completeness among predictors (Fig S17). In general, demographic and economic factors are well-observed, with some notable exceptions, such as air transportation (6.7% missing). Unfortunately, several potentially informative predictors suffer from poor reporting (*e*.*g*., influenza immunization coverage), and are excluded from our analysis.

### Geographic variation

The correlations shown in Figure 3F offer a valuable top-down view of global influenza epidemiology, yet these summaries mask salient variation among countries. For a more granular view, we also provide detailed country-level scatter plots of select outcome-predictor pairs. In Figure S15, we show how testing intensity varies with (A) total population (*ρ* = −0.55) and (B) *per capita* GDP (*ρ* = 0.69), but in very different ways (we also note the low correlation between these covariates, *ρ* = −0.3; see below). For example, Canada, the USA, and China all have high *per capita* testing intensity adjusted for population (Fig. S15A), while Canada’s surveillance intensity is higher than the USA when adjusting for GDP (Fig. S15B). More generally, we observe a remarkably wide range of testing intensities between countries of comparable GDPs, such as the 1:140 ratio between Indonesia and Bhutan (*per capita* GDP~ $10, 000) and the 1:170 ratio between Germany and Denmark (*per capita* GDP~ $50, 000). These results further highlight persistent global surveillance gaps, both in high-income countries as well as the populous countries of India, Indonesia, Pakistan, and Nigeria, which together include over 2 billion people while falling within the bottom 5% of global *per capita* influenza surveillance.

In Figure S16, we show country-level details of the strongest statistical predictors of four notable outcomes (color shows region): A) IAV seasonality versus absolute latitude (*ρ* = 0.39), B) IBV zero weeks versus population (*ρ* = −0.45), and C) IBV variation in peak timing (peak MD, *ρ* = −0.71). For IAV seasonality (A), tropical countries do not exhibit a clear latitudinal gradient, while the highest seasonality is seen in countries of intermediate latitude (*e*.*g*., Azerbaijan and Kyrgyzstan). For IBV zero weeks (B), we observe considerable variation among countries of similar populations constrained by an apparent upper bound in the most populous countries (*e*.*g*.,, India and Nigeria) and a lower bound in less populous countries (*e*.*g*.,, Qatar and Iceland). For IBV peak timing variation (C), we again find no clear latitudinal gradient among tropical countries, but we do find a sharp decrease towards less variable peak timing above approximately 40 degrees absolute latitude.

### Comparing IAV and IBV

As with temporal correlation (above), we find broad agreement of seasonal outcomes between these IAV and IBV. In Figure S14, we quantify the correlation structure among each pair of outcomes. Overall, we find that outcomes for IAV and IBV are highly correlated: *ρ* = 0.67 (positivity), *ρ* = 0.73 (seasonality), *ρ* = 0.41 (zero weeks), and *ρ* = 0.78 (inter-season variation in peak timing). In addition, we find a high correlation between seasonality and zero weeks for both IAV (*ρ* = 0.82) and IBV (*ρ* = 0.93).

In Figure 4, we compare IAV and IBV for select outcomes: positivity, B) seasonality, and C) variation in peak timing (color shows region). In A), IAV positivity is much higher than for IBV, though no regional pattern is apparent. In B), IAV seasonality is lower than for IBV. The regional pattern in C) is particularly evident, with most N Temp countries experiencing limited variation in peak timing while tropical countries experience much higher variation for both IAV and IBV.

**Fig. 4.**
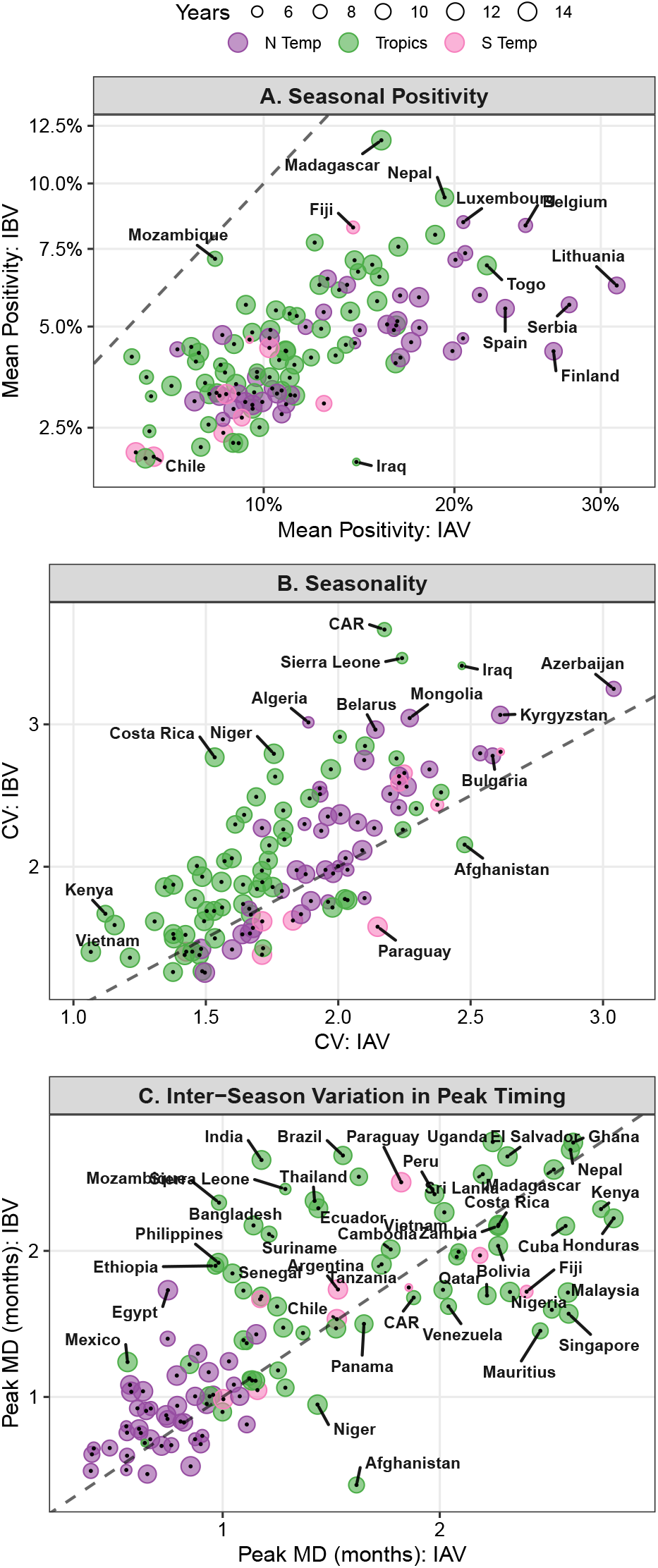
Comparison of select epidemiological outcomes (rows) between IAV (X) versus IBV (Y) grouped by region (color). In each case, we find a strong correlation between IAV and IBV: A) positivity (*ρ* = 0.67), B) seasonality (*ρ* = 0.73), and C) inter-season variation in peak timing (peak MD, *ρ* = 0.78). In A), positivity of IAV is much higher than for IBV, though no marked regional pattern is evident. The situation is reversed in B), where IAV seasonality is somewhat lower than for IBV. For peak MD in C), N Temp countries exhibit lower and more consistent variation in peak timing than tropical countries. Point size shows duration of record; dashed line shows 1-to-1 line.

## Discussion

In this work we have synthesized 15 seasons of global public health surveillance to reveal consistent geographic and socioeconomic patterns of influenza epidemiology. In contrast to prior studies (29), we find noteworthy congruence between IAV and IBV on a broad range of epidemiological outcomes, from temporal correlation (Fig. 2B) to seasonal outcomes such as positivity, seasonality, and fade-out dynamics (percent zero weeks) (Fig. 4 and Fig. S14). We document how fade-outs scale predictably with population size (among other factors), and demonstrate that IAV typically precedes IBV within a given season (Fig. 3F). We also identify strong associations between geography and seasonal timing, particularly between absolute latitude and inter-season variation in peak timing (Fig. 3G). Overall, our descriptive analysis demonstrates that while geographic and climatic factors remain critical drivers of influenza epidemiology, so too do socioeconomic and demographic factors. Integrating these factors into influenza modeling frameworks could enhance predictive accuracy, particularly in regions with under-resourced surveillance systems.

### The Human Geography of Influenza

An intriguing observation that emerges from our study is the occurrence of spatially clustered groups of countries with similar outcomes. In some instances, such as percent zero weeks (Fig. 3C), we identified mechanistically plausible predictors of the observed patterns, including population size and *per capita*air travel rates, consistent with metapopulation theory and the findings of phylogeographic models (18, 20). In other cases, our high-level view identified intriguing patterns that may have arisen from local or regional circumstances that were not immediately evident, such as a group of high-positivity countries in Western Europe, and the notably different peak IAV month in many Central and South American countries (Fig. 3A and 3D). Our results suggest that detailed attention to the particular circumstances of countries with related outcomes could uncover important factors driving influenza surveillance and transmission dynamics.

Another key finding of our work is wide-ranging differences in influenza epidemiology between N Temp and tropical countries, particularly regarding temporal correlation (Fig. 2C) as well as seasonality and inter-season variation in peak timing (Fig. 3). Furthermore, our clustering analysis (Fig. 2A and Fig. S8-S9) indicates that tropical countries comprise a complex mixture of different dynamics, with some countries behaving more or less similar to N Temp or S Temp countries. This heterogeneity has important consequences for public health efforts, many of which have been calibrated to the consistent seasonality of temperate countries (**?**), as well as potential ecological and evolutionary consequences as influenza lineages disperse between countries (51).

#### Climatic factors

Our findings underscore substantial latitudinal variability in influenza dynamics while revealing similarities in the spatial distribution of IAV and IBV. Consistent with established transmission patterns, we observe strong latitudinal gradients of seasonal intensity and timing (43–46). However, we also find significant variation in seasonal patterns across these gradients (*e*.*g*., Figs. S16A and S16C), suggesting that socioeconomic status, demographic composition, and health infrastructure also play a more influential role in shaping influenza dynamics than previously recognized (76, 77).

#### Demographics and socioeconomics

Our results underscore the central role of wealth in global public health. Furthermore, we note strong correlations among predictors of wealth and poverty, including *per capita* GDP, absolute latitude, *per capita* air travel, healthcare expenditure, and birth rate (Fig. S13). These widespread inter-dependencies underscore the value of a holistic approach when considering putative drivers of global influenza dynamics. Disentangling the relative impacts of climate, demographics, human mobility, and public health efforts (including testing and vaccination) on the influenza dynamics, particularly in tropical countries, represents a key research challenge.

#### Human Mobility

Future work that accounts for proximity and connectedness between countries and regions could shed additional light on our findings. In this work we have excluded a detailed analysis of connectivity due to the lack of such records for many LMICs. Nonetheless, our results show both broad patterns of temporal correlation by distance and region (Fig. 2C), as well as more granular details, such as the distinct timing of IAV seasonality found throughout Central and South America. We expect that many of these patterns reflect the influence of interregional human mobility, including air travel and migration, which can synchronize epidemics across geographically distinct regions(19, 78). The impact of human mobility on local elimination (i.e., percent zero weeks) remains an especially interesting avenue for future work, underscored by widespread interruptions in transmission observed during the SARS-CoV-2 pandemic.

### Surveillance Challenges for Global Public Health

A key objective of our study was to include as many countries (and seasons) as possible in order to provide a comprehensive picture of global influenza epidemiology. All told, we included observations on 111 of the world’s 195 countries, including all of the world’s top-10 most populous countries. However, our results also highlight persistent gaps in influenza surveillance, including low *per capita* surveillance in many of the world’s most populous countries (Fig. S15A) and a complete absence of routine surveillance throughout much of sub-Saharan Africa (Fig. 3), along with both periodic and sporadic gaps across the historical record (Figs. S1-S2).

These surveillance gaps exacerbate a frequent concern in epidemiology, that of distinguishing absence of evidence from evidence of absence. For example, we found that seasonal surveillance gaps were common in N countries (Fig. S2), presumably driven by local troughs of disease prevalence. However, these gaps interfere with reliable estimates of local extinction (e.g., zero weeks). Here we exclude weeks that lack surveillance data from our analysis, though we expect that these missing weeks are more likely when influenza is absent.

A related complication is the interplay between local disease prevalence and testing intensity, evinced by seasonal patterns of testing in N and S Temp (but not tropical) countries (Fig. S1). While we can not rule out the possibility that our results have been affected by the seasonality of testing, we did assess the relative timing of each season’s maximum of samples, IAV, and IBV, which we summarize in Figure S5. Here we find a close synchrony between testing and IAV: in 70% of country-seasons, these measures peak within 4 weeks of each other. The correspondence is much weaker for IBV (54% of country-seasons within 4 weeks), suggesting that testing is driven primarily by IAV and that some caution is warranted when interpreting IBV estimates of seasonality.

While subtype-level influenza analysis can offer valuable insights into influenza dynamics (16, 17, 41), our analysis does not include subtyping of IAV and IBV due to substantial gaps in lineage identification across countries and years. As shown in Figure S6, the *Not Determined* category consistently accounts for the largest proportion of IBV cases, and a similar lack of resolution is evident for IAV subtypes in many countries. This widespread missingness limits our ability to draw robust subtype-specific conclusions at the global scale. To reduce bias and ensure comparability across countries, we have focused here on aggregate IAV and IBV patterns rather than disaggregating by subtype.

Our analysis is also limited in some respects by our use of countries as a unit of observation. Countries are imperfect units of observation owing to A) each country’s composition of spatially distinct populations and to B) the wide variation among countries in geographic extent and population size. At the same time, each country has a unique cultural, demographic, and administrative history that presumably influences disease surveillance and control efforts. And, while more fine-grained analysis of influenza dynamics in, *e*.*g*., cities has provided valuable insights into the impacts of weather and other local factors (21, 23, 43), we note that no comparable, globally representative surveillance record for cities exists. With the above in mind, we focus here on large-scale patterns and long-term means, including climate and seasonality. We also note that our analysis of correlation by distance employs population-weighted centroids in order to better represent each country’s unique settlement patterns.

The SARS-CoV-2 pandemic presented a major shift in global public health that is clearly evident in our results. After falling sharply from 2019 to 2020, the median global *per capita* samples tested approximately doubled from its pre-pandemic baseline in 2023 and 2024 ((55, 56); Fig. S3). In the US, for example, samples tested jumped from 5.1 per 1,000 people in 2019 to 13.2 in 2024. However, these impacts were highly variable, and major interruptions in influenza surveillance have persisted, particularly across N Temp countries (Fig.S1). Meanwhile, pandemic controls on travel and other human contact led to stark global reductions in influenza case reports across much of 2020 and 2021 (Fig. S2). We do not directly quantify shifts across the pandemic here due to our country-level focus and the relatively few post-pandemic seasons currently available for analysis. Nonetheless, disentangling the overlapping impacts of shifting surveillance and transmission patterns across the pandemic era remains a central challenge to better understanding the pandemic’s role in shaping influenza epidemiology.

## Materials and Methods

### Data

#### Epidemiological Data

We obtained weekly country-level type-specific IAV and IBV data from WHO FluNet Interactive ILI and Viral Surveillance (79) for 182 countries total from 1995 to 2025 (coverage varied by country). We focus here on the 2010 to 2025 influenza seasons in order to minimize variation in the duration of surveillance among countries.

We label countries according to geographic region based on their WHO classification listed in the FluNet “FLUSEASON” field: NH=Northern Temperate (N Temp), SH=Southern Temperate (S Temp), and YR=Tropics. We note that the population centroid of most (but not all) countries categorized as Tropics is, indeed, within the geographical Tropics (absolute latitude *<* 23.5 degrees); notable exceptions including China and Bangladesh.

We define countries’ epidemic season according to their geographic region, with start months of May (N Temp), April (Tropics), and January (S Temp). For N Temp and S Temp regions, this corresponds to the month where most countries experience their (annual) minimum case count – that is, we choose the consensus trough month. Among Tropical countries, however, there is no consensus trough month and we instead choose the month where the fewest countries experience their maximum case count in order to avoid the peak month of the greatest number of countries.

We then employ the boundaries of the above epidemic seasons to compute uniform criteria for excluding countries and seasons that lack sufficient data to estimate seasonality. We excluded country-seasons with A) fewer than nine months (and at least one week per month) or 30 weeks of non-missing IAV case reports, B) fewer tests than IAV cases (mostly countries that did not report samples tested), and C) fewer than 20 IAV cases. Due to low IBV reporting in many countries and seasons, we do not employ exclusion criteria for IBV cases. We do, however, exclude countries with a 2010 population size of less than 100,000.

#### Geographic and Socioeconomic Data

Annual country-level data on demographic, economic, social, climatic and geographic factors were obtained from publicly available sources, including the World Bank Data Portals (80–83), the Organization for Economic Cooperation and Development (OECD) (84) and WHO (85). Table S.1 provides a detailed overview of each factor. Coverage varies by country, and we exclude predictors with less than 90% coverage of included country-seasons.

The effective location of each country was determined using a population-weighted centroid rather than a geographic centroid to better reflect each country’s unique settlement patterns. We used the 2015 gridded population count data from WorldPop Hub (1km UN adjusted) (86) to compute this centroid for each country. Population centroids were then used to A) compute distance between country pairs and B) to compute the absolute latitude of each country.

Temperature, precipitation, and relative humidity were available on a monthly basis, and were averaged across each country’s respective epidemic season. All other indicators were available annually. Percent of country area classified as arid and tropical were computed from World Bank maps of Köppen-Geiger Climate Classification (1976-2000).

Time-invariant factors include absolute latitude, percent arid, and percent tropical. For the remaining factors, we compute the median value across the included epidemic seasons for each country. We note that variation in each factor is generally much higher among countries than among years within a country, motivating our use of a single representative value for each country. We also exclude several factors from further consideration due to their high overall correlation with another factor (*ρ >* 0.9, shown in parentheses): percent of population age 0 to 14 (with birth rate), per capita health care expenditure (GDP), and WGI’s government corruption index (government effectiveness). See Figure S13 for additional details.

### Statistical Analysis

#### Software

Statistical analysis was conducted in R 4.5 (87). We used the `mgcv` package for constructing generalized additive models (GAMs) (88) (`gam(meothd=“REML”, …)`), and the `ggeffects` package for computing estimated marginal means (89).

### Temporal Correlation

This analysis was conducted separately for IAV and IBV. We first computed the pairwise Spearman correlation (*ρ*) between all pairs of countries, and then grouped country pairs by region (e.g., both countries in Tropics) or in different regions. We then constructed a mixed effects generalized additive model (GAM) that predicted *rho* from (categorical) country-pair region and distance (smooth term), along with country identities as random effects. For each GAM, we exclude country-pairs with fewer than 2 years (104 weeks) of overlapping data, and weight observations by the square root of observed weeks.

For comparison, we also assess distance metrics (geographic centroid and minimum boundary distance) and find broadly comparable results (Fig.S10).

### Seasonal epidemiology

This analysis tabulated a set of seasonal outcomes by country from weekly case reports of IAV and IBV (10 for each type; see Figure 3). Outcomes included: per capita samples tested (i.e., sampling intensity), positivity, and percent zero weeks (no reported cases). Within each season, seasonality was computed from the coefficient of variation (CV) of weekly case reports. We also identified the month of peak total cases, and computed the inter-seasonal variation in peak timing as the maximum absolute deviation (MD) of peak month using circular arithmetic. Finally, we tabulated the relative difference in peak timing between IAV and IBV.

We then constructed a set of statistical linear models using country and season as random effects. We have one model per response and type except for testing and relative peak timing (no type) and variation in peak timing, where only one observation per country was available. We use a negative binomial GLM for the proportional responses with an appropriate offset: testing intensity with log(population) and positivity with log(samples tested). We use a binomial GLM for percent zero weeks (i.e., where each week constitutes a binomial trial) and a standard linear model for seasonality.

For each model, we computed the estimated marginal mean response for each country (R function `ggeffects::predict-response(margin=‘empirical’, …)`).

This procedure allowed us to account for systematic variation among epidemic years to estimate a single mean response per country. We note that, similar to covariate data, average differences among countries of each response are much greater than year-to-year variation within each country. This observation, along with different observation periods across countries, motivated our choice to summarize outcomes by country rather than country-season. Consequently, each country is weighted equally in the subsequent analysis (rather than weighting by observation period).

Using these mean responses, we computed the Spearman correlation (*ρ*) of the mean response with each covariate (Figure 3G). We also compute the *ρ* between each pair of responses (Figure S14) and covariates (Figure S13). We then adjust p-values (two-sided test for difference from zero) using a false discovery rate correction with *α* = 0.05 (66).

## Data Availability

All data was obtained from public sources. All data and code will be made available at osf.io upon publication.

