## Supporting Figures for "Global comparison of influenza A and B epidemiology identifies consistent geographic and socio-demographic predictors"

March 13, 2026

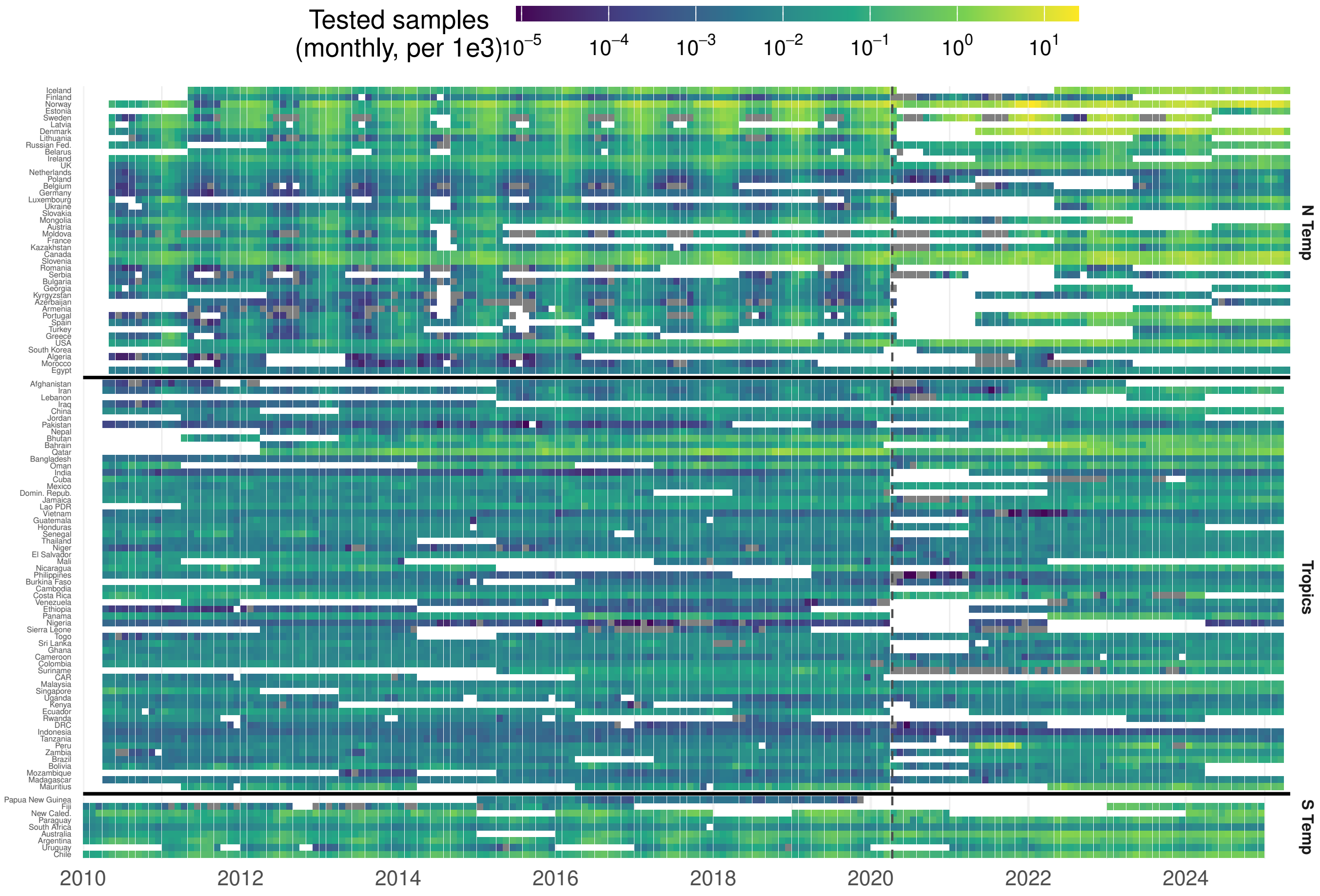


Figure S1: Monthly per captia samples tested (log scale). Grey shows zeros; white shows missing data.

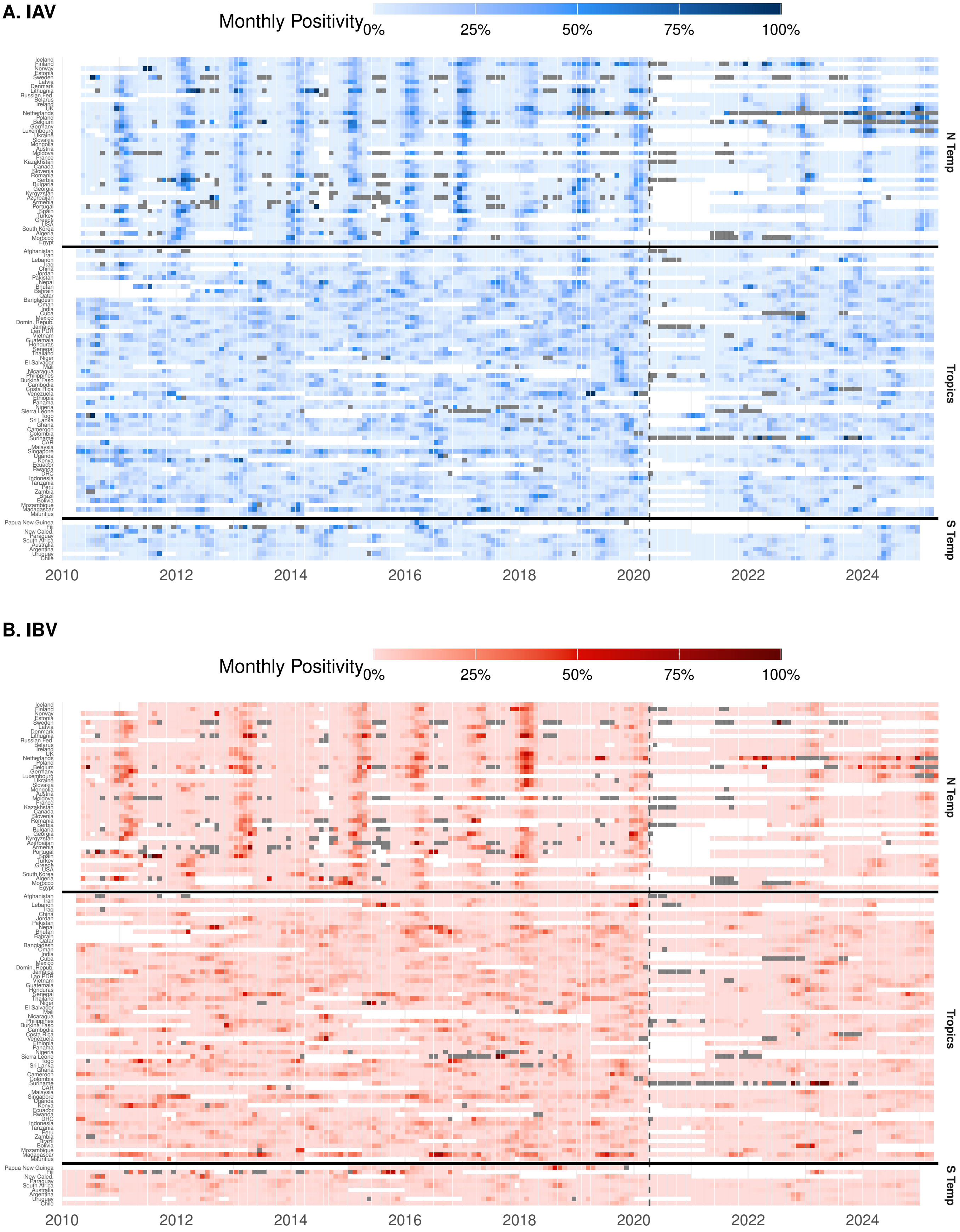


Figure S2: Monthly positivity (cases/samples) for A) IAV and B) IBV. Grey shows zero samples tested; white shows missing data. Horizontal black lines separate the three regions: North Temperate, Tropics, and South Temperate. Within each region, countries are ordered from north to south. Vertical dashed line shows beginning of the 2020 SARS-Cov-2 pandemic.

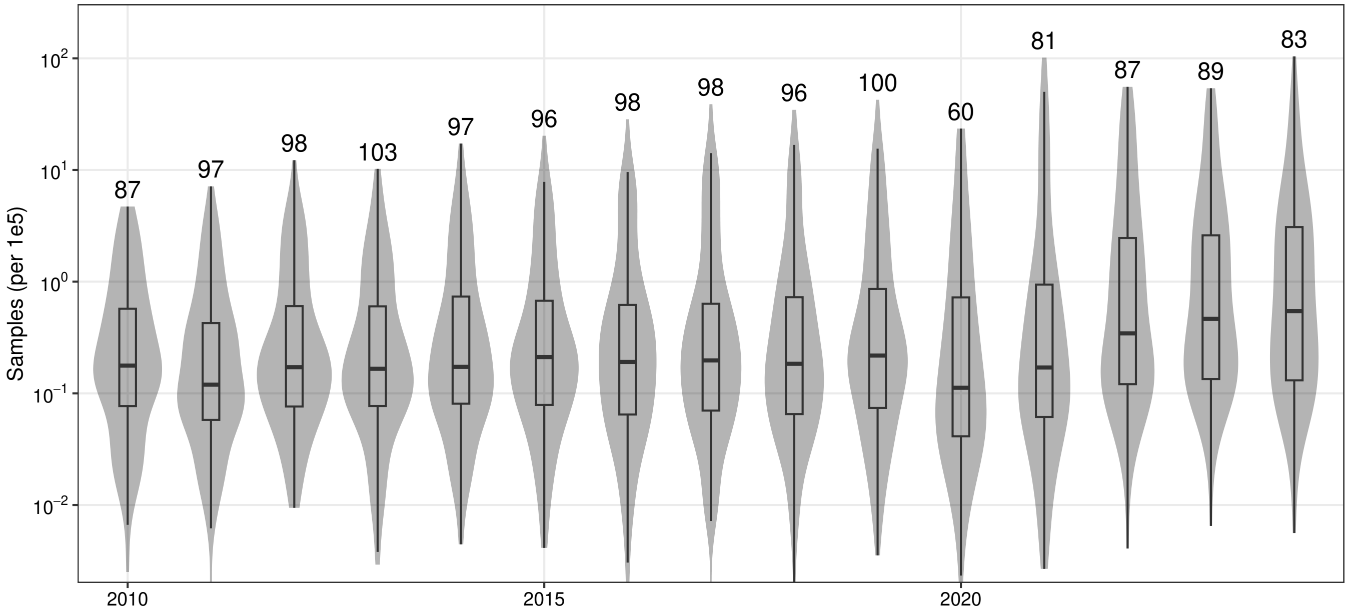


Figure S3: Yearly distribution of per-country samples tested. Box shows median and IQR; numbers show reporting countries per year.

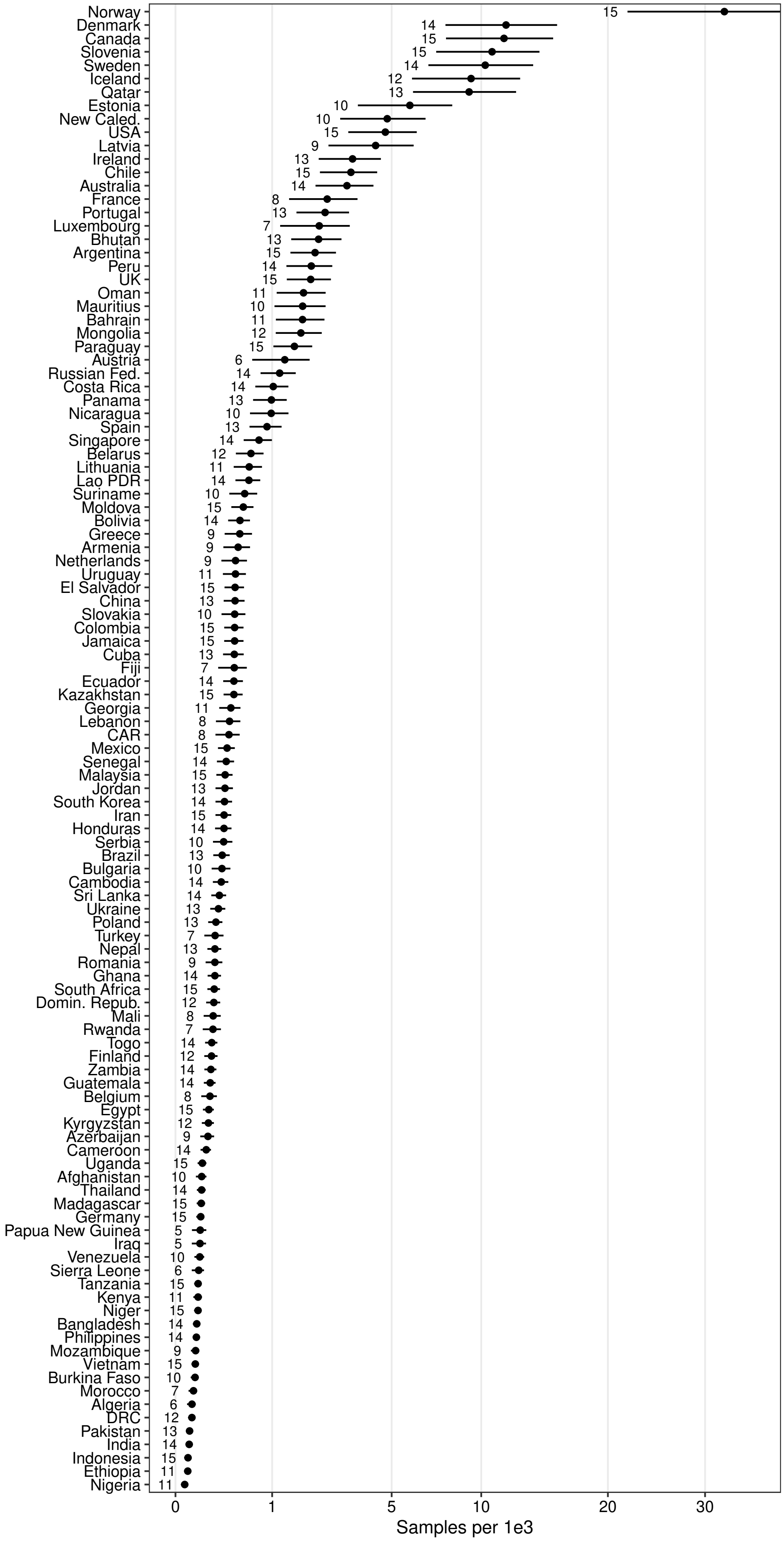

Figure S4: Per capita samples tested per season by country, showing estimated marginal means (EMM) +95% CI. Numbers show available seasons per country (2010-2015).

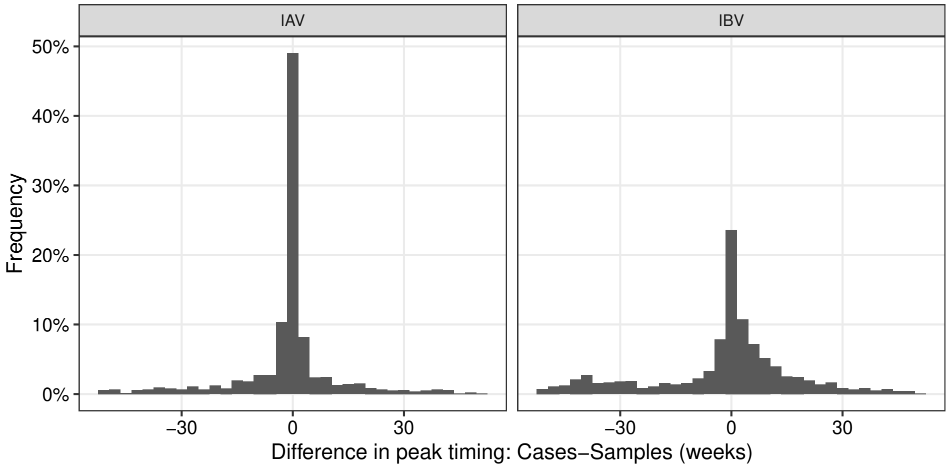


Figure S5: Relative timing of seasonal maximum of weekly samples tested versus case reports for IAV and IBV across all countries. Consistent synchrony between sampling and IAV cases is evident, with peaks in 70% of country-seasons within 4 weeks (versus 54% for IBV).

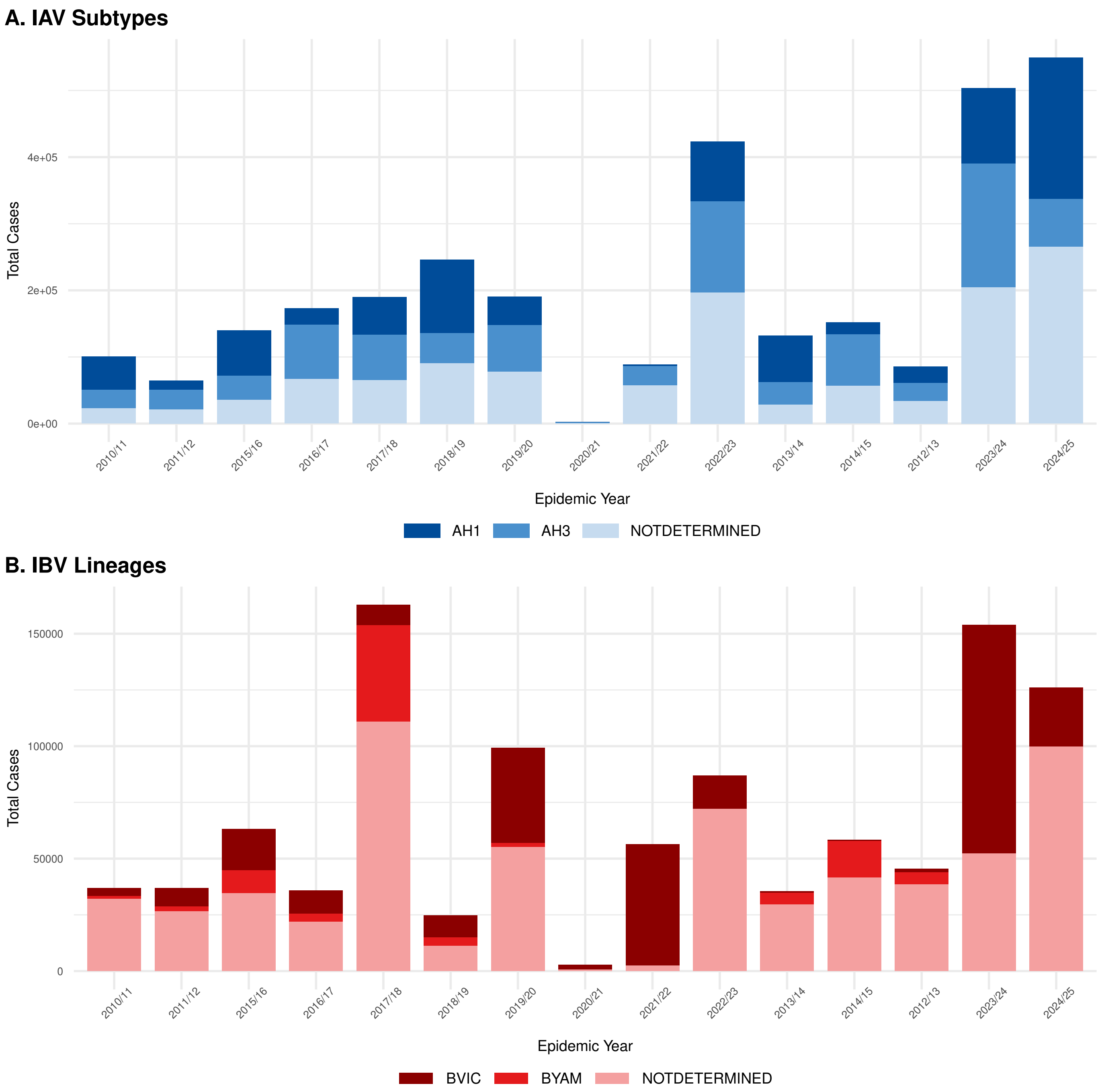


Figure S6: Case reports stratified by influenza subtype (when available). (A) Total reported cases of Influenza A subtypes (AH1, AH3, and A not determined). (B) Total reported cases of Influenza B lineages (Victoria and Yamagata) over the same period (n=898 rows). ?? An overall rise in surveillance is evident, yet as substantial portion of cases are not resolved to the subtype level in most years.

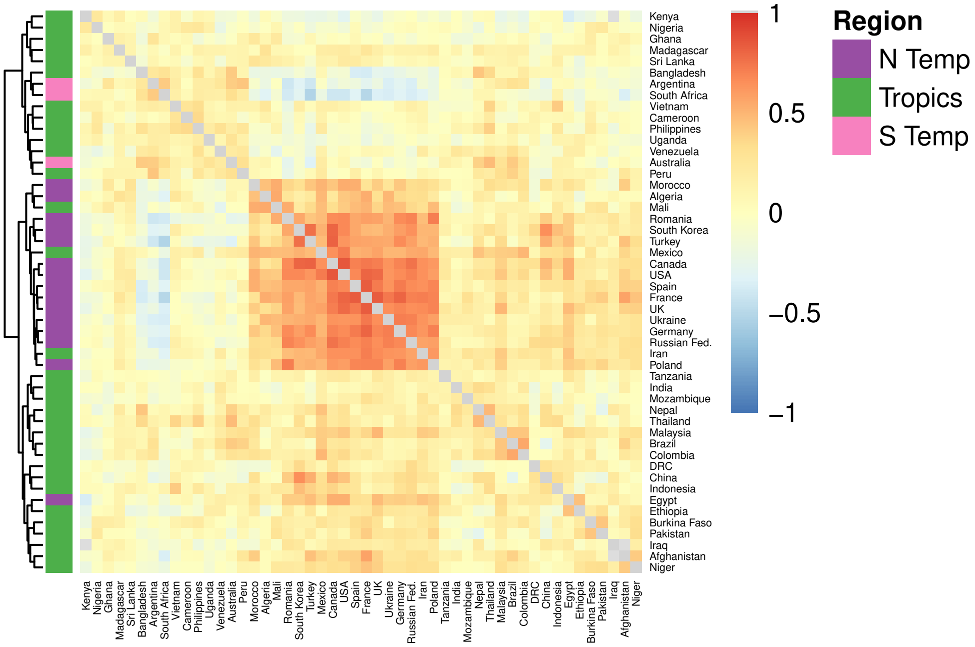


Figure S7: IBV temporal correlation, select countries. See also Figure 2.

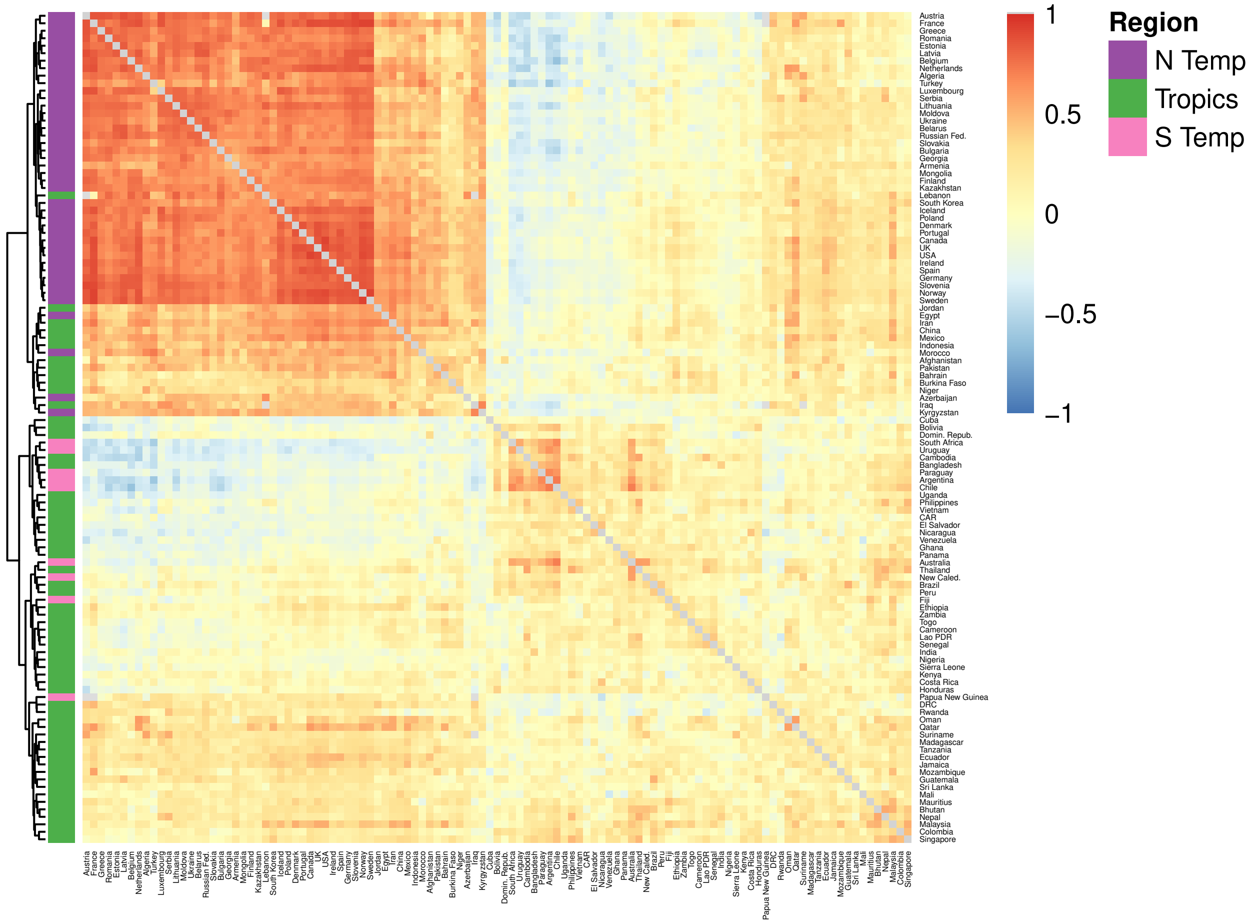


Figure S8: Temporal correlation: IAV, all countries. See also Figure 2.

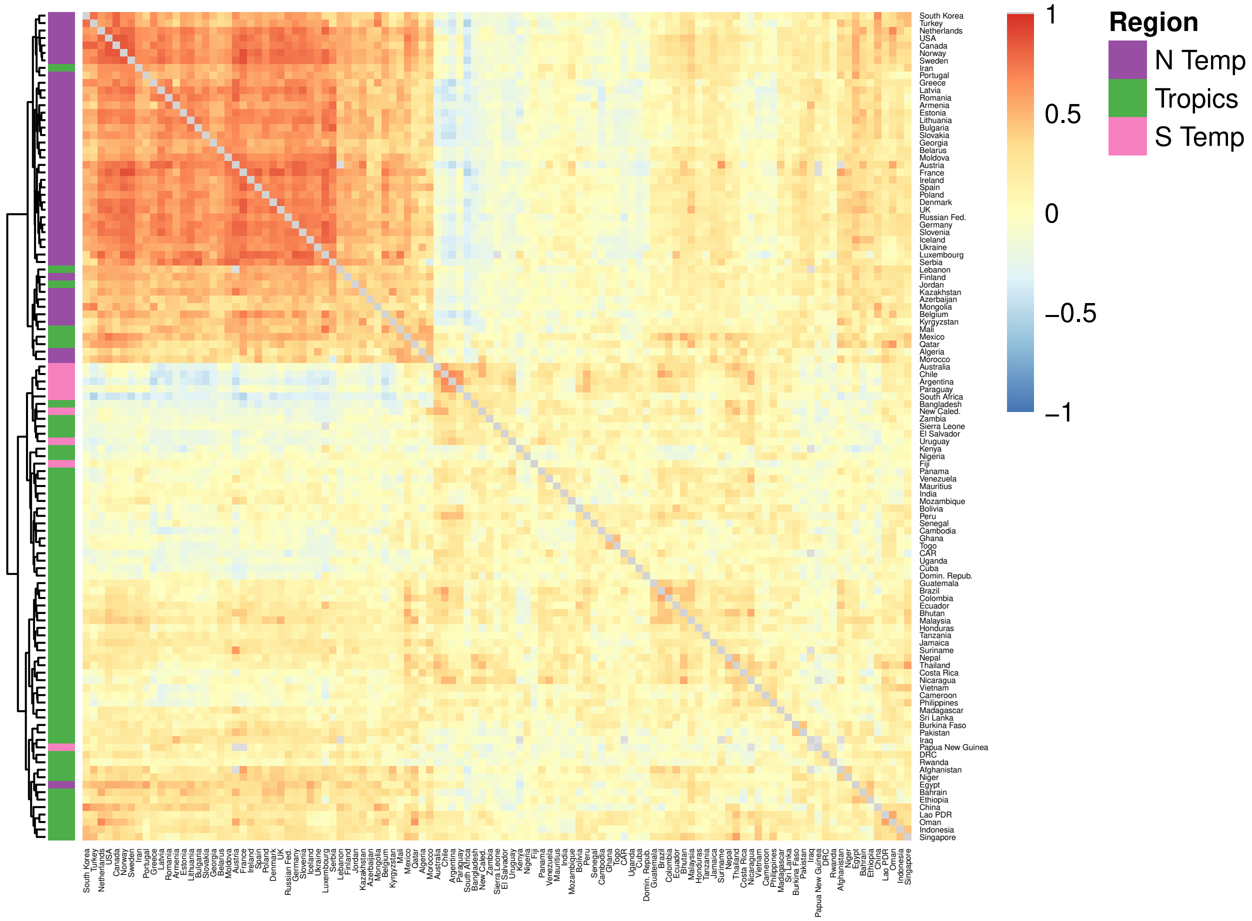


Figure S9: IBV temporal correlation, all countries. See also Figure 2.

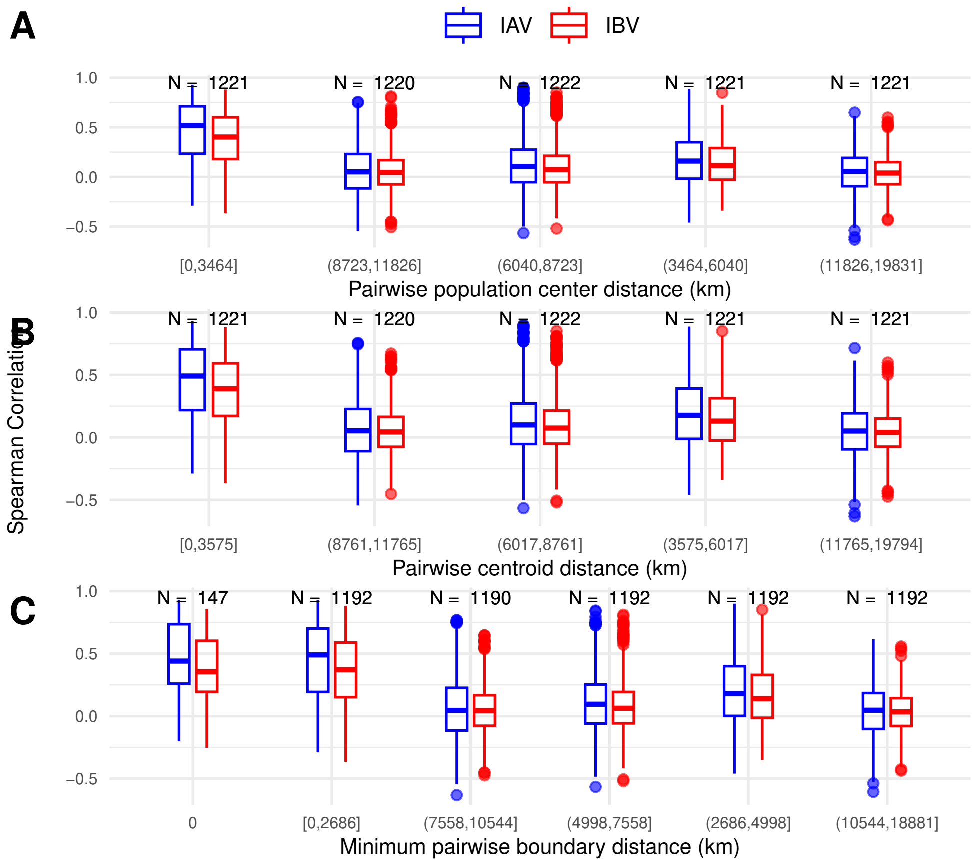


Figure S10: The distribution temporal correlation of weekly IAV and IBV cases between country pairs (Y, Spearman) versus distance (X) for A) population centroid distance, B) geographic centroid distance, and C) minimum boundary distance.

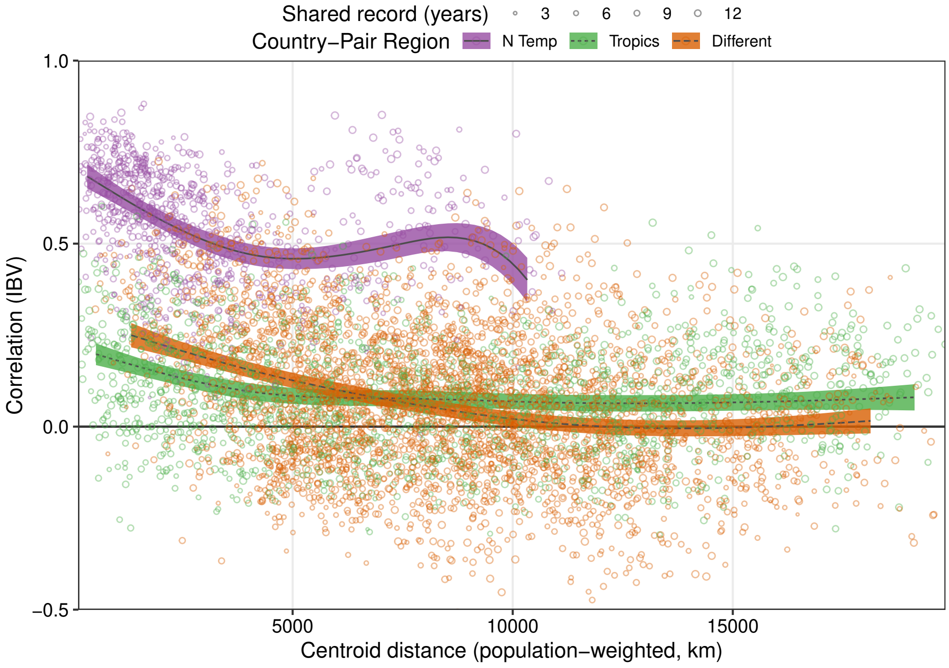


Figure S11: Model results for IBV showing estimated correlation ($\rho$) by distance (X) and region (color), as in Figure 2C.

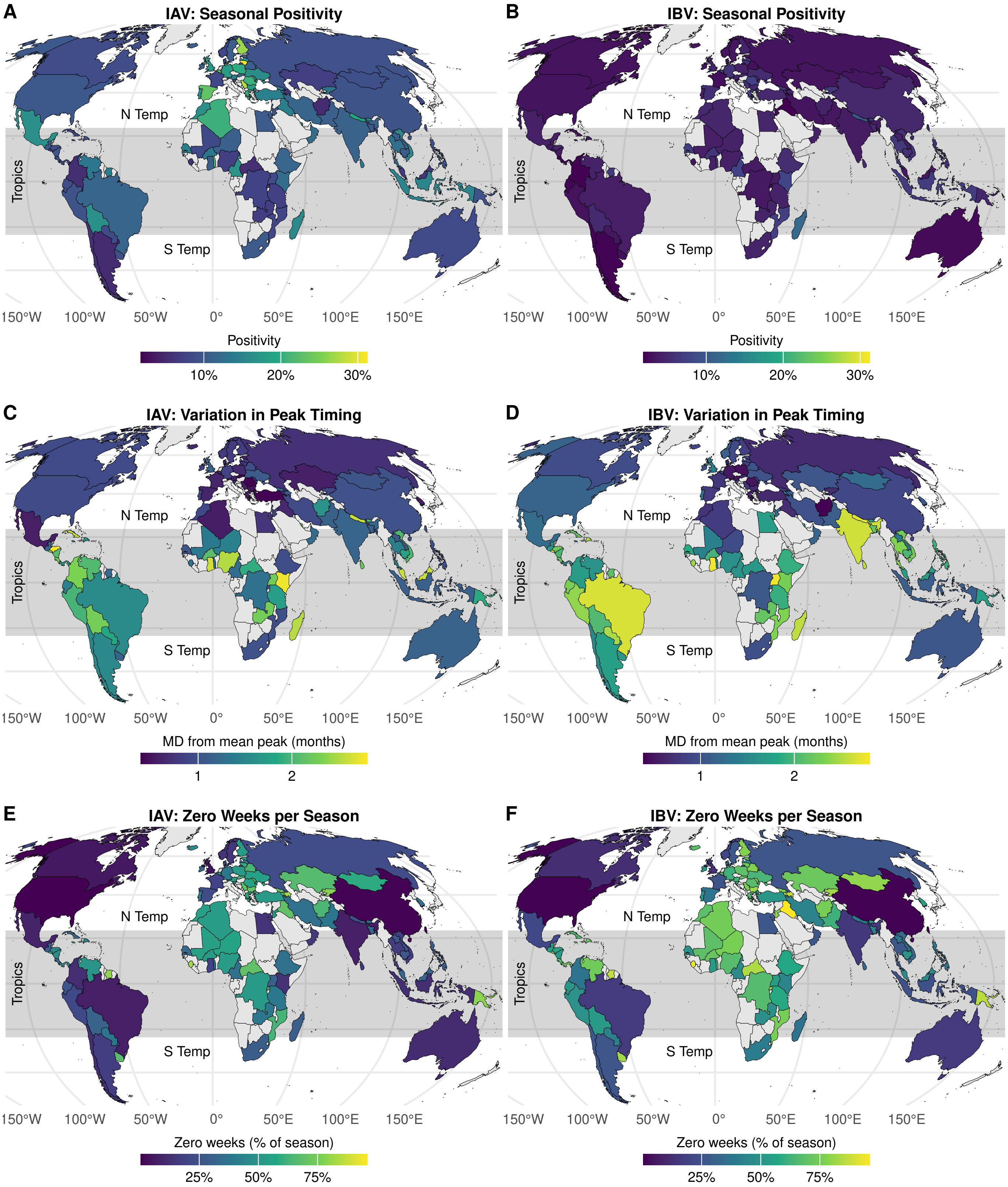


Figure S12: Geographic comparison of IAV vs IBV (columns) of three epidemiological outcomes (rows): A) positivity, B) peak MD, and C) entropy. Overall, IAV positivity is much higher than IBV (A), while a much stronger latitudinal gradient is apparent for peak MD (B) and entropy (C). See also Figure 3 and 4.

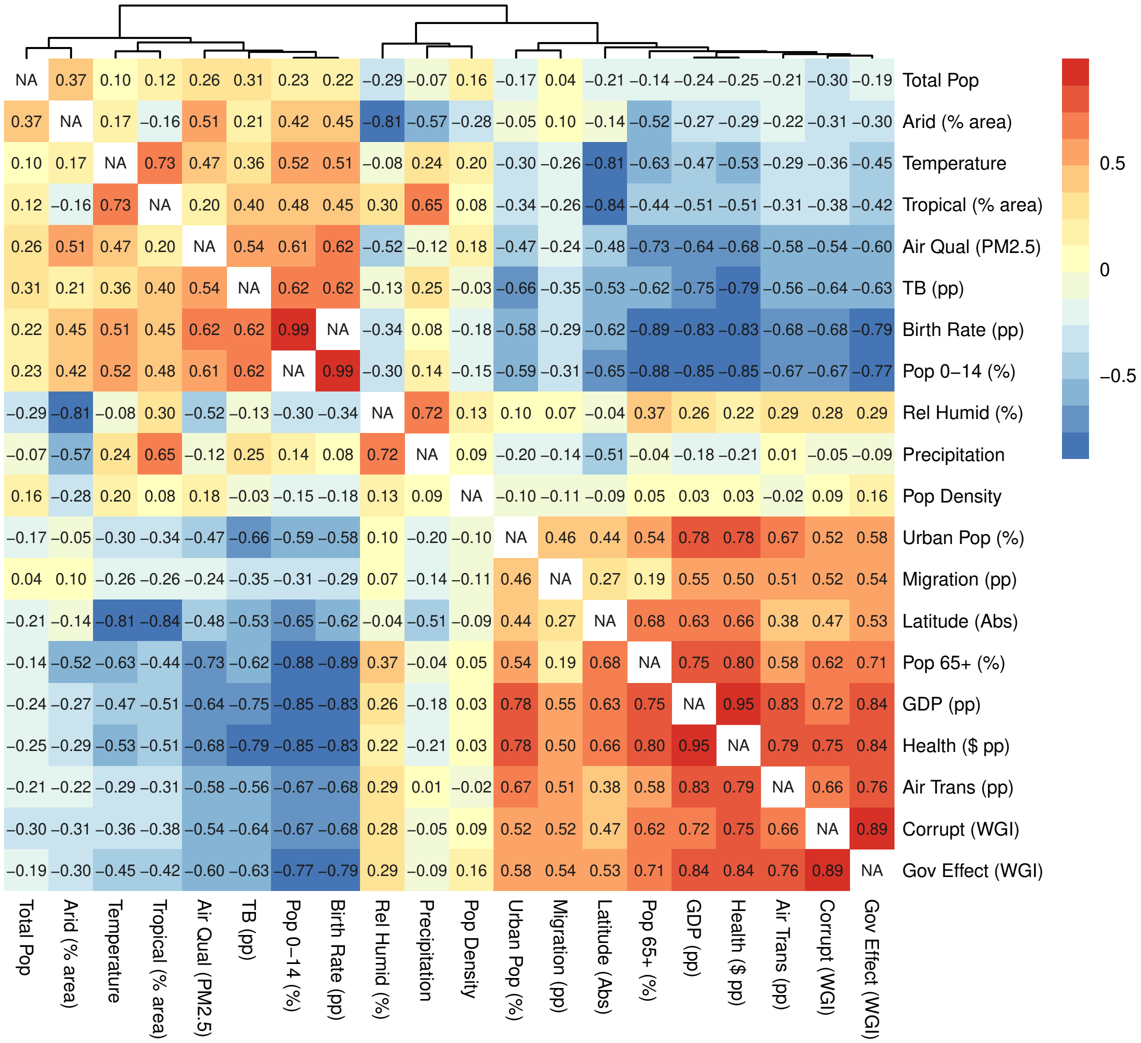


Figure S13: Spearman correlation ($\rho$) between each pair of putative predictors (country medians). See also Figure 3.

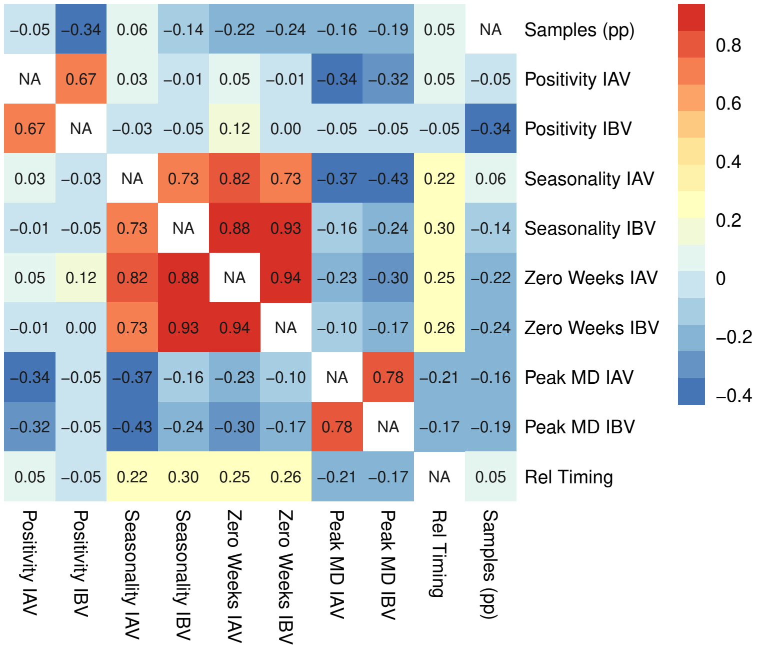


Figure S14: Spearman correlation ($\rho$) between each pair of seasonal epidemiological outcomes. See also Figure 3.

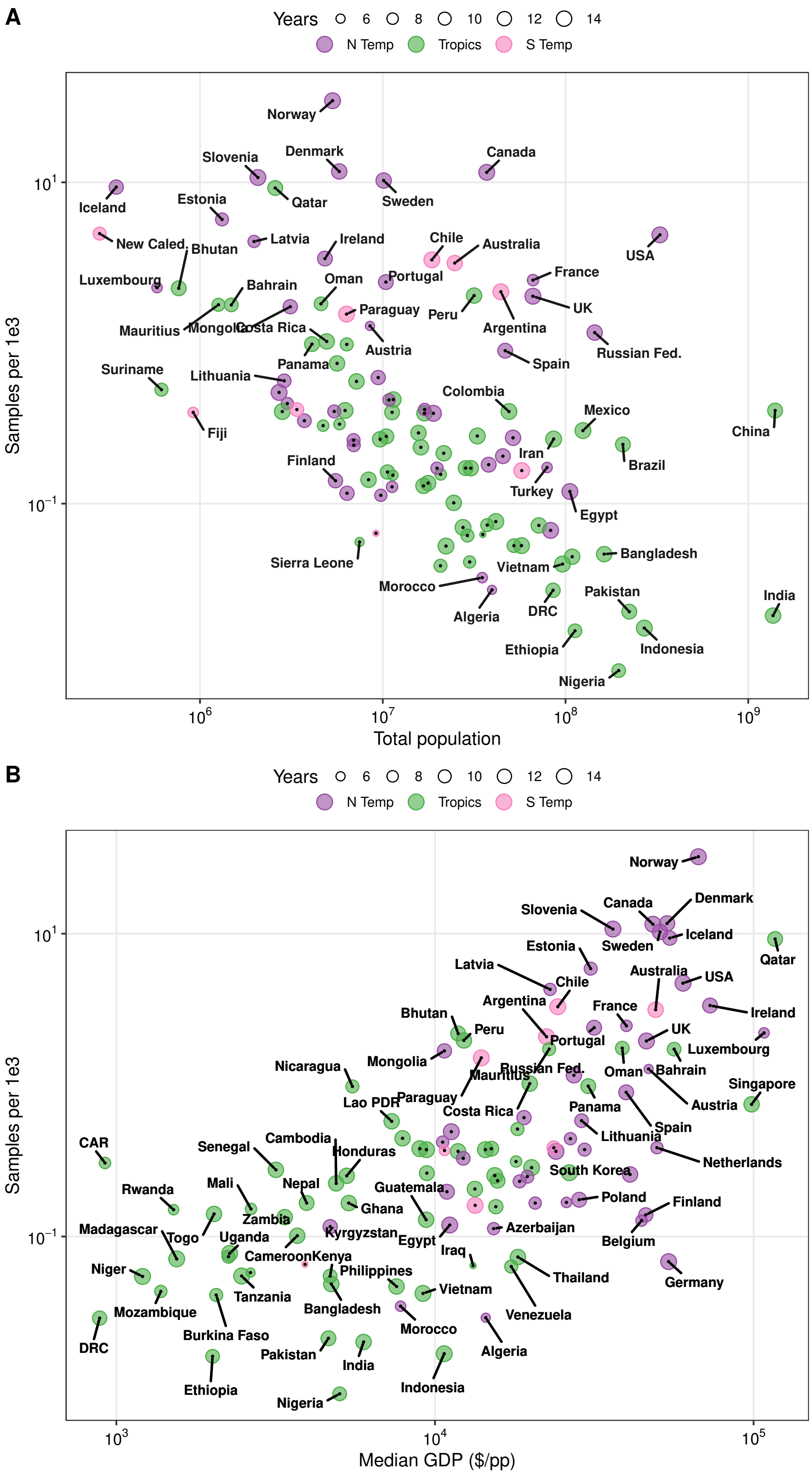


Figure S15: Noteworthy statistical predictors of samples tested per capita : A) total population ($\rho = -0.55$) and B) median GPD ($\rho = 0.69$). See also Figure 3.

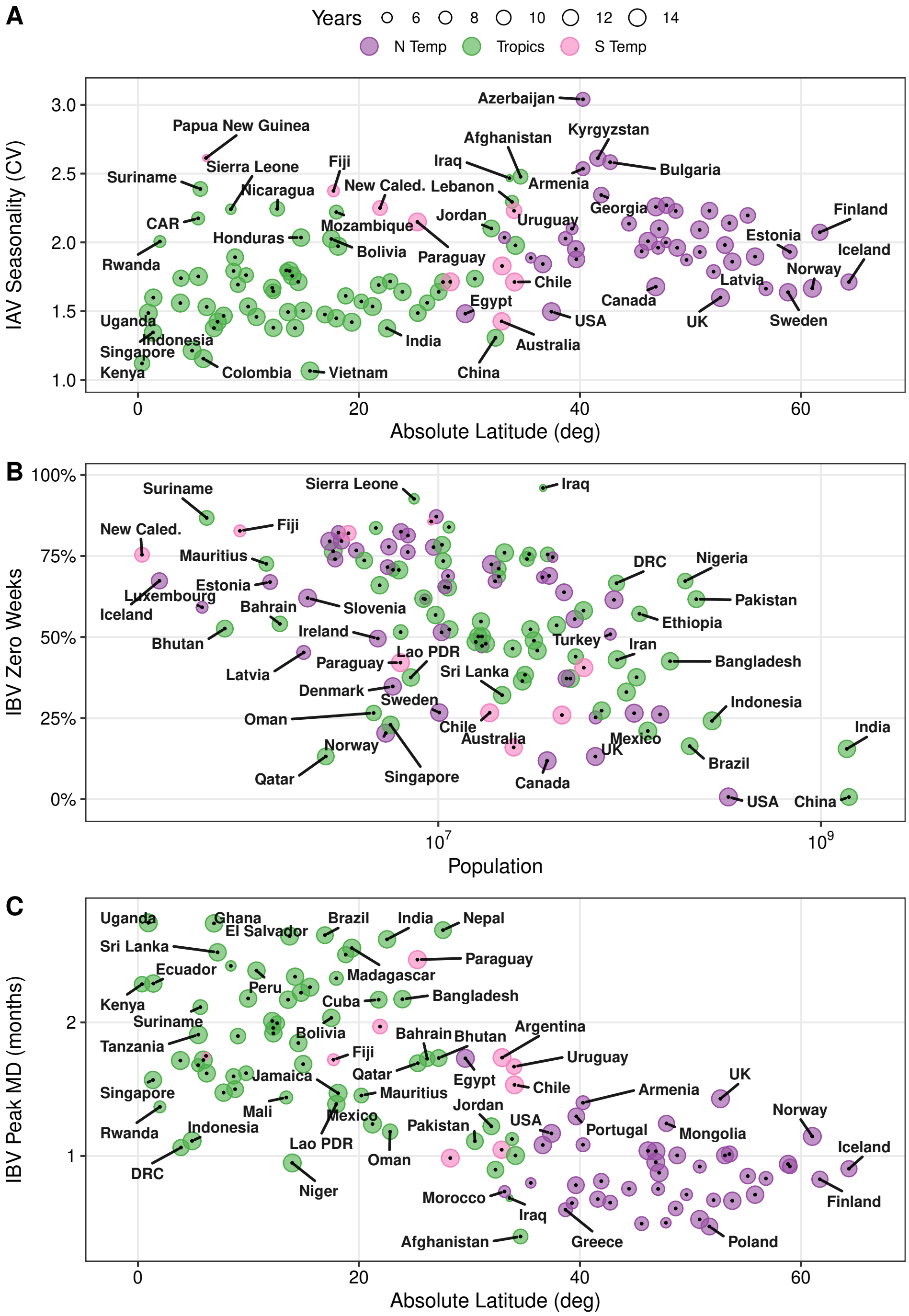


Figure S16: Country details of select epidemiological outcomes (Y) paired with their strongest statistical predictors (X) and grouped by region (color): A) IAV seasonal intensity versus absolute latitude ($\rho = 0.39$), B) IBV zero weeks versus population ($\rho = -0.47$), and C) IBV inter-season variation in peak timing versus absolute latitude ($\rho = -0.71$). Point size shows duration of record. See also Figure 3.

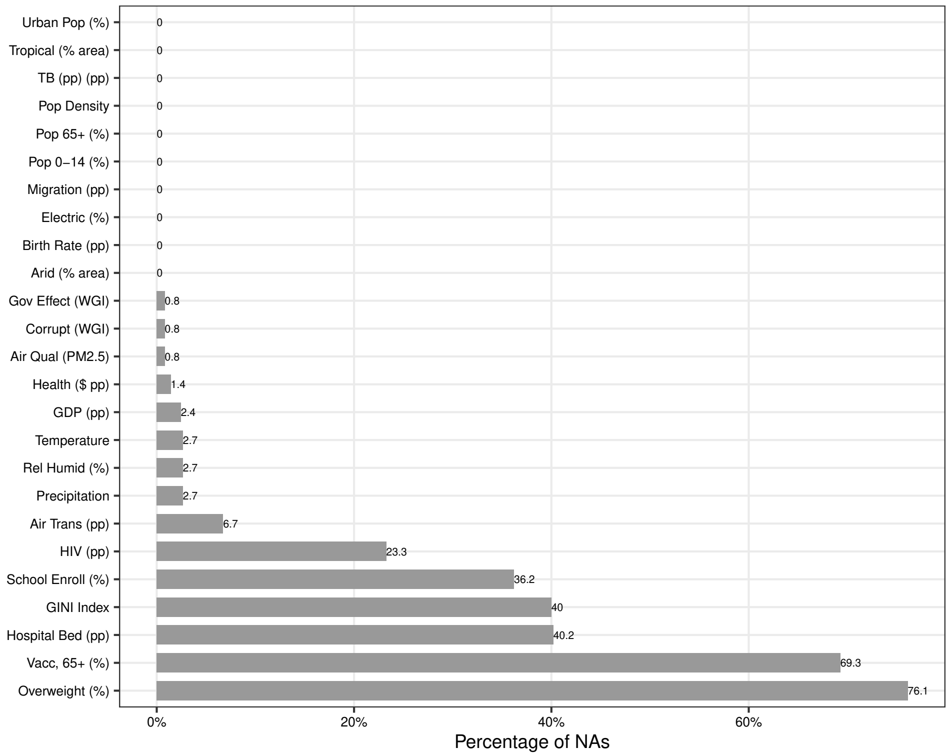


Figure S17: Frequency of missing data for each predictor.
